# COVID-19 Observations from Hospitalized Ward Patients in the Northern Emirates: A Practice Only Preached

**DOI:** 10.1101/2021.10.20.21265254

**Authors:** Shahab Qureshi, Drishti D. Kampani, Tara Ali Hassan Al-Qutbi, Aalya Mohamed, Mubarak Alfaresi

**Affiliations:** Sheikh Khalifa General Hospital, Umm Al Quwain, United Arab Emirates; University of Sharjah, Sharjah, United Arab Emirates

## Abstract

**Background:** The COVID-19 pandemic has established itself as the defining global health crisis of this time. The study describes the clinical profile of hospitalized, non-ICU patients with COVID-19 in the United Arab Emirates (UAE) during its second wave, through January-March 2021. It also highlights the use of antibiotic stewardship principles in patients admitted with COVID-19.

**Methodology:** An observational, retrospective study was conducted with 110 participants from Sheikh Khalifa General Hospital – Umm Al Quwain in the UAE. Pregnant women, patients who were admitted to/transferred to/discharged from the intensive care unit, patients who were receiving antibiotics prior to admission were excluded from the study.

**Results:** Population was 58.2% male with a mean age of 51.2 (± 14.6) years; 69.1% had at least one comorbidity and 61.8% were classified as severe COVID-19 disease. Mean WBC count was 6.03 ± 2.70 × 10^9^ cells/L with a mean CRP of 83.3 ± 14.6 mg/L. 4.2% of the tested (20.9%) blood cultures performed were positive. Immunomodulators (67.26%), prophylactic anticoagulants (90%), anti-viral drugs (83.61%) were primary modalities of therapy. Empiric antibiotic use was limited to 9.1% of population.

**Conclusion:** Our study highlighted that the population admitted to the hospital in the second wave of the COVID-19 pandemic in the UAE were mostly male, older with higher prevalence of comorbidities. Given the limited knowledge of the new disease, we took bold but calculated clinical measures to maintain antibiotic stewardship practice and brought antibiotic prescribing to extraordinary low level not seen during the COVID-19 pandemic.

## Introduction

The COVID-19 pandemic has firmly established itself as the defining global health crisis of this time and one of the greatest challenges that we continue to face as a global community. First discovered in December 2019, the world has come a long way since - from battling a novel, unknown virus to rolling out international vaccination programs which are working to bring down the morbidity and mortality associated with the disease.

Globally, there is a sharp contrast between COVID-19 status with infection rate rising in in many countries with few of them still at the peak of their infection curve while the other countries are displaying a downward trend. (1) Infection rates have largely been disproportionate – with some populations affected worse than the others due to a host of reasons ranging from high medical comorbidity profiles to socioeconomic challenges. (2) The absence of a ‘one-size fits all’ strategy makes it essential to learn from the experiences of different centers around the world. Understanding which strategies worked and which strategies have the potential to work from a clinical perspective contribute to advancing global knowledge and pandemic preparedness.

In the United Arab Emirates, COVID-19 infections have sharply reduced with an average of 304 new infections reported each day, at 8% of its peak (as of 29^th^ September 2021). (3) These figures are in comparison to the peak noted on 31^st^ January 2021. UAE experienced a sharp increase in the number of cases, colloquially called the ‘second wave’, during the periods from January to March 2021. (4)

Prior to this, the initial increase of COVID-19 infection rates or the ‘first wave’ was seen from March to August 2020. In the United States, 80% of deaths associated with COVID-19 were among adults aged ≥ 65 years with the highest percentage of severe outcomes among persons aged ≥ 85 years. In contrast, a recent study published from the first COVID-19 center in the UAE, patients were young (mean age ± SD = 49 ± 15 years) with diabetes and/or hypertension and associated with severe infection as shown by various clinical and laboratory data necessitating intensive care unit (ICU) admission. (5)

During the months of January to March 2021, the UAE imposed several restrictions across different sectors – tourism, business, entertainment, and healthcare. (3, 6) Rapid immunization programs were accelerated by making four different COVID-19 vaccines available, free and on an opt-in basis: Sinopharm, Pfizer-BioNTech, Sputnik V, and Oxford-AstraZeneca. In March 2021, seven COVID-19 field hospitals were opened across the country. (7) However, in the early phases of the increase, when these field hospitals were still not functional, majority of the case load was borne by the government hospitals who were overwhelmed with the number of cases. Admission criteria for COVID-19 was revised to only admit symptomatic patients who required clinical monitoring due to limited bed availability.

Furthermore, in clinically deteriorating patients with viral infections such as those admitted during the second wave, a major concern is usually a secondary or co-infection with bacteria contributing to increased morbidity and mortality. A similar belief about COVID-19 was widely prevalent. (8) Is it prudent to use antibiotics empirically in non-ICU hospitalized COVID-19 patients? Through this case series, the study also hopes to elaborate on its experience on reducing empiric antibiotic use in COVID-19 patients in Umm Al-Quwain.

To our knowledge, there has been no formal publication that describes the clinical profile of hospitalized, non-ICU patients with COVID-19 in the UAE during the second wave in 2021. This study, conducted in Sheikh Khalifa General Hospital- Umm Al-Quwain, aims to retrospectively review the clinical profile of these patients and highlight the use of antibiotic stewardship principles in patients with COVID-19 in Umm-Al-Quwain.

## Methodology

### Study population

This clinical observational study included symptomatic confirmed COVID-19 patients admitted to general wards in Sheikh Khalifa General Hospital - Umm Al Quwain (SKGH-UAQ), United Arab Emirates between 15^th^ January to 31^st^ March 2021. SKGH-UAQ is a federal hospital operated by the Ministry of Presidential Affairs, serving the population of the Umm Al Quwain emirate and adjacent northern emirates of UAE.

Patients who are 20 years old or above and were diagnosed with COVID-19 through a confirmed positive RT-PCR test were included. Pregnant women, patients who were admitted to/transferred to/discharged from the intensive care unit (ICU), patients who were receiving antibiotics prior to admission were excluded from the study.

### Data Collection

After review by the Research Ethics Committee of University of Sharjah (REC 21-07-15-01-S) and approval from the hospital board, data was collected from the written and electronic health records of included patients within the hospital.

Sample size calculation with a confidence level of 95% and margin of error of 5% for UAE’s population of 9.89 million in 2020, (9) determined the minimum required sample size to be 384 participants. However, this study includes a total of 128 patients. Every subject meeting the criteria of inclusion has been selected within the specified time frame of January to March 2021. The time frame cannot be extended to include the calculated sample size as the variables of the study are representative of the ‘second wave’ of COVID-19 and are therefore, time-sensitive.

Data covered the following sections – (i) demographic data (age, gender); (ii) pre-existing chronic co-morbidities such as diabetes mellitus (DM), hypertension, cardiovascular disease (CVD), chronic lung disease, chronic kidney disease (CKD), stroke/transient ischemic attack (TIA), cancer, and any other documented comorbidities; (iii) radiological features on chest x-ray and/or high-resolution computed tomography (HRCT) scan; (iv) COVID-19 disease severity (mild, moderate, severe, critical); (v) laboratory investigations (white blood cell count, platelet count, C-reactive protein, ferritin, D-dimer, blood culture, respiratory panel); (vi) pharmacological regimens (immunomodulators, anti-coagulants, anti-viral drugs, supplements, empiric antibiotics); (vii) duration of stay in the hospital.

Disease severity was classified based on National Institute of Health COVID-19 Treatment Guidelines: (a) mild disease, if patients have any signs or symptoms of COVID-19 without dyspnea, shortness of breath or abnormal chest imaging; (b) moderate disease, if patients have evidence of lower respiratory disease by clinical assessment or imaging and oxygen saturation above 94% on room air at sea level; (c) severe disease, if respiratory rate ≥ 30/min, blood oxygen saturation (SpO2) < 94%, the fraction of inspired oxygen (FiO2)] < 300, and/or lung infiltrates > 50% within 24 to 48 h; (d) critical disease, if patient has respiratory failure, septic shock and/or organ failure. (10)

The laboratory information collected have the following reference values: (a) white blood cell count (4.0-10.0 × 10^9^ cells/L), (b) platelet count (150 – 450 × 10^9^/L), (c) c-reactive protein (0-10.0 mg/L), (d) ferritin (15-200 μg/L), (e) d-dimer (0 – 0.49 μg/mL). Respiratory pathogen panel was performed whenever possible, with the primary goal of ruling out influenza infections.

### Statistical analysis

Data was imported from Microsoft Excel to IBM SPSS, version 25 (IBM Corp., Armonk, NY, USA), for analysis and interpretation. After applying the exclusion criteria, data for 110 participants was analyzed.

For continuous variables, data has been presented as mean and standard deviation. Categorical variables have been represented using frequencies and percentages. Data was visualized using Canva Chart Designer (Canva®) to prepare the figures.

## Results

A total of 110 participants were included in the study. Out of these participants, 58.2% and 41.8% identified as males and females respectively, with a mean age of 51.2 (± 14.6, range = 20 – 89) years.

The majority of the patients (69.1%) had at least one comorbidity, prior to diagnosis of COVID-19. The type and percentage of patients with each comorbidity is represented in Figure 1

**Figure 1.**
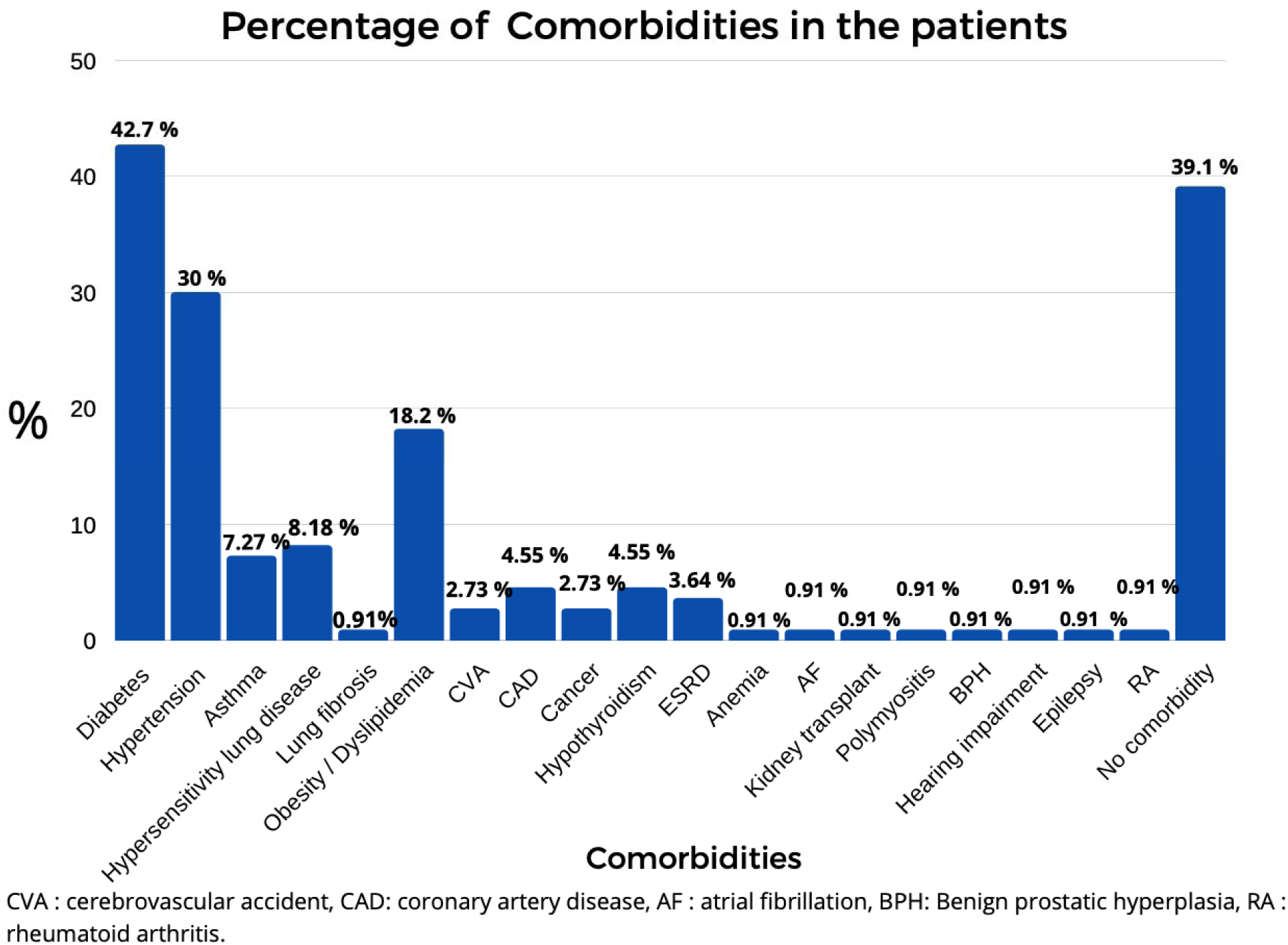
graphically represents the distribution of comorbidities in the population of patients admitted with COVID-19 in the general ward of SKGH-UAQ, UAE between January-March 2021.

White blood cell count, platelet count, c-reactive protein, ferritin and d-dimer levels were assessed in admitted patients as prognostic factors, recommended under the UAE Ministry of Health and Prevention (MOHAP) guidelines. (11)

Blood culture was performed in 20.9% of the patients, of which 95.6% showed no growth, while 4.2% had blood culture positive for Staphylococcus hemolyticus. The investigation was not performed for the remaining 79.1% of the patients; decision made on clinical assessment. Co-infection with influenza was tested for 52.8 % patients and was reported as negative in all of them.

Chest X-ray was performed for all patients with the following findings – increased broncho-vascular markings (21.8%), bilateral opacities (65.5%), unilateral lobe opacity (1.8%) and no appreciable disease (10.9%). High resolution computed tomography (HRCT) was performed only in 5.5% patients, showing diffuse ground-glass opacities in these patients.

Three lines of pharmacotherapy were used to manage the patients in our wards – a) immunomodulators, b) prophylactic anticoagulants, and c) anti-viral drugs as elaborated in Table 1.2. Empiric antibiotics were used for very few patients as elaborated in Figure 2.

**Table 1.1.** illustrates the mean value of laboratory investigations undertaken for patients admitted with COVID-19 in the general ward of SKGH-UAQ, UAE between January-March 2021. It highlights the distribution of the population within and above the reference range.

| Lab Investigation | WBC | Platelets | C-reactive protein | Ferritin | D-dimer |
| --- | --- | --- | --- | --- | --- |
| **Reference range** | 4.0 - 10.0 x 10<sup>9</sup> cells/L | 150 – 450 x 10<sup>9</sup>/L | 0-10.0 mg/L | 15-200 µg/L | 0 – 0.49 µg/mL |
| Mean value | 6.03 ± 2.70 | 217 ± 90.69 | 83.3 ± 14.6 | 763 ± 68.25 | 0.97 ± 0.64 |
| % Patients <u>within</u> the reference range | 71.8 % | 78.2 % | 10.9 % | 24.8 % | 25.5 % |
| % Patients <u>above</u> the Reference range | 7.27 % | 2.73 % | 89.1 % | 75.2 % | 74.5 % |

**Table 1.2.**
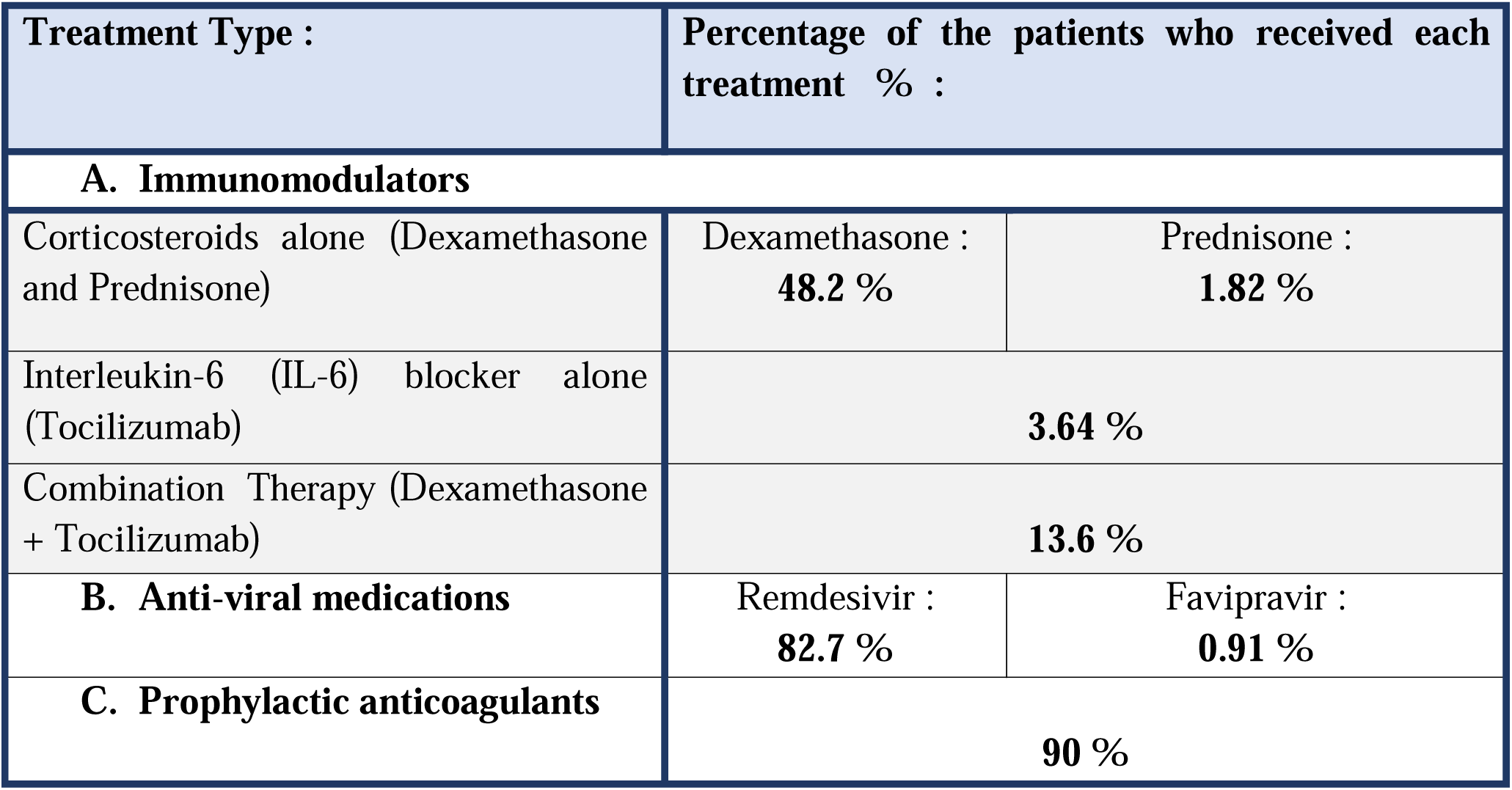
outlines part of the pharmacotherapy used to manage COVID-19 in patients admitted to the ward between January-March 2021, based on UAE MOHAP guidelines.

**Figure 2a.**
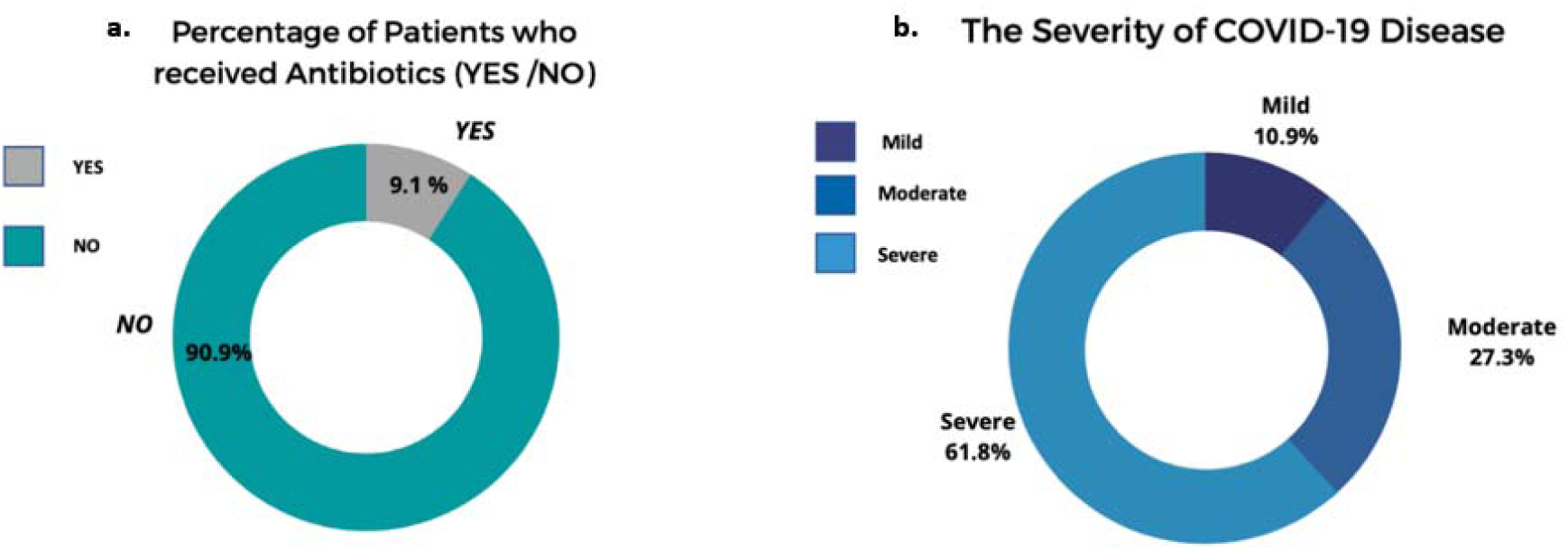
highlights the limited use of empiric antibiotic therapy in patients admitted to the ward. Figure 2b. represents the spectrum of COVID-19 disease in the wards based on the UAE MOHAP guidelines.

In summary, the patient’s clinical and radiological assessment determined whether the patient’s disease was classified as mild, moderate, and severe. Figure 2 demonstrates the distribution of patients, further highlighting that majority of the patients had severe COVID-19 disease.

The preliminary outcome of the study was to determine mortality on the ward during the duration of the study. 100% of the patients included in the study, who received treatment in the general ward were discharged alive. As a secondary outcome, duration of stay was assessed in the ward. The mean duration (in days) of stay from presentation to discharge was 6.2 (± 3.8 days, range = 0 - 25) days, with a mode of 3 days.

## Discussion

The COVID-19 pandemic ushered in a healthcare crisis that left behind more questions than the ones it answered. In such uncertain times, experiences from single centers like the Sheikh Khalifa General Hospital, Umm Al Quwain in UAE serve as important pieces in the large puzzle. Our study highlighted that the population admitted to the hospital in the second wave of the COVID-19 pandemic in the UAE were mostly male, older with higher prevalence of comorbidities compared to the patient profile noted in the first wave of the pandemic in the UAE.

The patients admitted to the ward had failed outpatient care and were rapidly worsening; they were severely sick patients requiring oxygen. We followed guidelines for treatment as per UAE’s Ministry of Health and Prevention (MOHAP) updates and treated majority of our hypoxic patients with close observation. (11) Less than 5 percent of our patients were moved to ICU due to requirement of high flow nasal cannula (HFNC) to maintain oxygen levels >94%.

Our protocol for treatment with immunomodulators and antivirals were based on MOHAP guidelines and also took into account international consensus such as Infectious Diseases Society of America (IDSA) and National Institutes of Health (NIH) protocols. 90% of our patients also received anticoagulants as recommended by American Society of Hematology (ASH) and International Society on Thrombosis and Hemostasis (ISTH).

We based our use of antibiotics on our initial and daily assessments. We clinically assessed our patients for signs and symptoms of bacterial superinfection. Our assessment included the following: (a) cough with sputum production, (b) change in sputum color over the past 48 hours, (c) white cell count with neutrophilia, and (d) evidence of dense lobar consolidation on radiography. Our success with minimal use of antibiotics was made possible by daily close observation, a well-informed close team and daily examination of our patients.

Since the onset of the pandemic, there has been increasing debate about the use of empiric antibiotics to mitigate the risk of morbidity and mortality associated with bacterial co-infections in patients with viral respiratory infections. This led to the use of empirical antibiotics in patients with COVID-19 through the pandemic. In a recently published systematic review, mean rate of antibiotic use in COVID-19 patients globally was in 74.0% of cases. However, only 17.6% of patients who received antibiotics had secondary infections and pooled data of 4 studies highlighted that more than 50% of patients receiving antibiotics were neither severe nor critical. (12) Furthermore, another review study revealed that the overall proportion of COVID-19 patients with bacterial infection is merely 6.5%, with majority in those who were critically ill. (13) Through 2020, we noticed increasing analyses of bacterial co-infections showing an average of 3% community-acquired bacterial superinfection to as high as 14% hospital-acquired superinfection. (14). A regional COVID-19 study also showed similar infection rate data. (15)

Prior to the COVID-19 pandemic, higher c-reactive protein levels have traditionally been considered to be associated with a bacterial infection source. (16) The value of this practice has been called into question in our set of patients. The elevated levels of CRP might be linked to the overproduction of inflammatory cytokines in severe patients with COVID□19. (17) Thus, CRP production is induced by inflammatory cytokines and by tissue destruction in patients with COVID□19. From the perspective of the clinical manifestations, the essence of COVID-19 should be viewed as a sepsis induced by viral infection. (17)

The viral sepsis induced by SARS-CoV-2 has the typical pathophysiological characteristics of sepsis, that is, the early cytokine storm and the subsequent immunosuppressive stage. Upon SARS-CoV-2 virus challenge, alveolar macrophages and epithelial cells release a large number of cytokines and chemokines. (18, 19) Monocytes and neutrophils may be recruited to the site of infection and clear the exudate containing virus particles and infected cells, in turn leading to uncontrolled inflammatory response. During this process, adaptive immunity is difficult to start effectively due to the significant decrease of lymphocyte number and T-cell mediated immune dysfunction. Therefore, a deep understanding of the clinical significance of CRP in the diagnosis, treatment, and prognosis of sepsis in SARS-CoV-2 infection is helpful to the early rationale in using antibiotics (20).

The limitation of this study lies in its relatively small sample size. Combined with its design as a single-center study, there is limited generalizability of the results. Lack of information on post-discharge health status of these patients could be a potential limitation addressed by future studies to expand knowledge on management of this illness.

There is growing concern for a potential rise in antimicrobial resistance due to increased antibiotic prescription for COVID-19 patients around the world. The potential of bold strategies as adopted in our study, pave the way forward to cement existing antimicrobial stewardship programs. A recent study conducted in the UAE identified that 63.4% medical professionals follow national health authorities as their primary source of COVID-19 information. (21) Thus, a national framework would serve as a context-specific reference covering education about antimicrobial resistance, tracking local patterns, development of clinical guidelines and antibiotic control.

## Conclusion

The second wave of the COVID-19 pandemic ushered in a difficult time with challenging patients for the United Arab Emirates. Our study is the first formal publication to describe the clinical profile of hospitalized non-ICU patients during the country’s second wave – they were male, older and had more comorbidities. Given the limited knowledge of the new disease, we took bold but calculated clinical measures to maintain antibiotic stewardship practice and brought antibiotic prescribing to an extraordinarily low level previously not seen during the COVID-19 pandemic. We hope to share our experience with the world and create an impactful discussion.

## Data Availability

All data produced in the present study are available upon reasonable request to the authors

## Acknowledgements

We would like to acknowledge the REC committee of University of Sharjah as well as Dr. Zuhair Afaneh, Chief Medical Officer of Sheikh Khalifa General Hospital – Umm Al Quwain for their support in facilitating the study.

## Author contributions

S.Q. conceived the study. S.Q., D.D.K., T.A.H.A. were involved in designing the methodology for the research, collection, and curation of data. D.D.K. performed the formal analysis and T.A.H.A. was responsible for data visualization. S.Q., D.D.K., T.A.H.A. wrote the original draft of the manuscript. A.M. and M.A. performed the internal review prior to submission.

